# Formative research in designing a health, nutrition, and child development digital intervention for urban refugees and host communities in Nairobi

**DOI:** 10.1101/2025.08.01.25332720

**Authors:** Joyce Marangu, Amina Abubakar, Nathaniel Jensen, Simbarashe Sibanda, Mark Tomlinson

## Abstract

**Background:** Despite advances in addressing child health and undernutrition, low- and middle-income countries in Africa have experienced a persistently high number of children with suboptimal outcomes in the last decade. Caregivers of young children in low-resource settings have limited access to health and nutrition information to enhance child development. Digital interventions are a possible solution, although factors for their success among urban, low-income refugee and host communities are unclear. This study sought to explore the perceptions of healthcare workers and caregivers on health, nutrition, and early child development practices; identify barriers and facilitators for suitable digital interventions; and apply findings to enable intervention design.

**Methods:** We conducted a formative study to inform an intervention design using a qualitative approach. This involved key informant interviews (n=12) and focus group discussions (2; n=17) with caregivers of children below two years of age. The key informants included health, nutrition and child development officials from the Ministry of Health, and representatives from community-based organizations. Data were analysed thematically.

**Results:** While there were some perceived positive practices among refugee and host communities, they were described by officials as being insufficient, especially around breastfeeding, dietary diversity, ante- and post-natal care and positive parenting. There were limited digital interventions despite increased access to smart phones. Barriers such as access to smart phones and digital literacy were seen as likely to hinder uptake of digital interventions. However, use of existing resources such as community health promoters were seen as facilitators of intervention uptake. The findings from the formative study were mapped to intervention decision making.

**Conclusion:** The findings from this study indicate potential for digital solutions. They also make an important contribution to designing effective, contextually appropriate, and sustainable digital interventions which have a higher likelihood of being accepted within the community.

## Background

Globally, maternal mortality rates have plateaued following a rapid drop between 2000 and 2015 from 227 to 223 per 100,000 live births. Low- and middle-income countries (LMICs) in Africa accounted for more than 90% of global births (1), and 70% of maternal deaths in 2020 (2). The high maternal mortality rates are attributed not just to obstetric causes, but also to social determinants such as challenges in seeking and receiving care, high poverty rates, low education levels among women, and conflict resulting in displacement.

Child mortality rates in LMICs remain high, at 74 deaths per 1000 live births, in contrast to 5 deaths per 1000 live births in high-income countries. Moreover, reduction in child malnutrition rates has been slow. Global stunting and wasting stands at 22% (148.1 million) and 6.8% (45 million), respectively. This represents an average decline of about 0.5 and 0.09 percentage points in stunting and wasting,respectively, between 2000 and 2022 (3). This indicates a need for more intensified efforts in maternal and child health and nutrition programs to meet the Sustainable Development Goals (4).

By 2050 it is estimated that over 60% of the global population will be urban (5). A quarter of the global urban population currently lives in informal settlements, with the highest numbers in Africa and Asia (6). Informal settlements often have poor housing, inadequate infrastructure, and limited access to basic services, including healthcare, water, and sanitation. Informality also increases vulnerability to climate disasters such as flooding and heat stress (7).

The number of refugees and displaced persons is at an all-time high due to conflict, violence, disasters and effects of climate change (8). The sub-Saharan Africa region remains both the largest source, as well as destination, for refugees (8). With greater economic prospects in cities compared to refugee camps, Nairobi, where this study takes place, has experienced a steady inflow of migrants and refugees from neighbouring countries in the Horn of Africa and Great Lakes region (9). While local policies confine refugees to camps situated in Northern Kenya, many refugees have found their way to urban areas and live in urban host communities. The host community (10) provides support, resources, and to some degree integration opportunities. Local integration is a favourable solution when repatriation is unsafe, as it ensures that refugees can rebuild their lives, contribute to the economy and support their children’s development.

Refugees living in urban informal settlements experience multiple challenges, including language barriers, lack of legal documentation and frequent movement disrupting continuity in care. These factors may contribute to poor health outcomes (11). Poverty among both refugees and host communities contributes to low health-seeking behaviour and affects health outcomes (12). Further, indicators for child health, including mortality, malnutrition, and childhood illnesses, are worse in urban informal settlements compared to rural households in Kenya (13-15).

Some effort has gone into addressing these challenges through interventions such as universal health coverage (16), Linda Mama free maternity programme (17), baby friendly initiative (18), affordable housing (19), and social cash transfers (20), all of which have varying levels of success. Digital technology presents a possible alternative in enhancing maternal and child health. Reviews on the effectiveness of mobile health (mHealth) technology in enhancing pregnancy outcomes have mixed findings (21-23). For instance, health promotion messaging and postnatal appointment alerts were found to have a positive effect on the utilization of antenatal care services, and reduction of maternal deaths (24). mHealth interventions were also linked to increased health seeking behaviour and vaccination rates (25), increased rates of kangaroo care, exclusive breastfeeding and appropriate complementary feeding (26), and enhanced health worker supervision (27, 28). However, effectiveness of mHealth interventions may be limited by low acceptance among healthcare providers, delayed response in crises, one-way non-interactive communication, and may not be suitable for complex behaviour change interventions (29).

Community perspectives, when developing an intervention, are critical for success. Perspectives should be sought not just from the end users, but also those who will deliver the intervention (30). This approach has been shown to be beneficial in shaping the focus to ensure its relevance to the community (31), and to increase cultural relevance (32), acceptability, feasibility and sustainability (33).

There is limited evidence on the inclusion of urban refugee and host communities in understanding practices in health, nutrition and child development for purposes of designing relevant digital interventions.

The objectives of the current study were: 1) to explore the perceptions of healthcare workers and caregivers of young children on current health, nutrition, and early child development practices among urban refugee and host communities; 2) identify barriers and facilitators for digital interventions; and 3) apply the findings to intervention design.

## Methods

### Study design

This study was formative research (34) to inform the design of an mHealth intervention seeking to enhance the quality of maternal and child data and outcomes through a smart phone application. It employed a qualitative approach, comprising focus group discussions (FGDs) and key informant interviews (KIIs), to explore the perspectives of healthcare workers and caregivers of young children.

### Setting

The study was conducted in urban informal settlements in Eastleigh, Kamukunji sub-county, Nairobi. With more than 268,276 inhabitants, Kamukunji is characterized by a high-density population of more than 25,000 people per square kilometre (35). In addition to the Kenyan locals, this population also comprises embedded refugees from neighbouring countries, among them Somalia, South Sudan, Ethiopia, and Uganda. Settling of refugees in Eastleigh is attributed to political instability in the Horn of Africa from as early as the 1980s (36). Eastleigh has experienced a strain on critical services, including water and sewage, health, education and security due to under-investment in these sectors (36). In common with other informal settlements in Nairobi (37), the population is characterised by high child malnutrition rates (38), which are linked to developmental delays (39).

### Sampling

Participants for both the focus group discussions and key informant interviews were selected through purposive sampling. The sample size (n=29) was determined by data saturation (40). Contacts for the key informants (n=12) were obtained through the Kamukunji Sub-County Ministry of Health, upon approval from the Nairobi County Directorate of Health. The sample comprised healthcare professionals and representatives of relevant community-based organizations within Kamukunji Sub-County.

Focus group discussion participants (n=17) were caregivers of children below three years, recruited from public health facilities and Community Based Organizations (CBOs) operating in the study site. There was a total of two focus groups, the first with eight and the second with nine participants, with a mix of both refugee and host communities. Four community health workers based in Eastleigh were engaged to mobilize the caregivers based on their knowledge of the community members and their previous work with refugees.

### Procedures

Semi-structured interview guides were developed after review of existing literature in line with the research objectives, dialogue within the research team, and pre-testing with a small sample of participants to enhance clarity of the questions and prompts. Field notes were used for early identification of themes and guided further refinement of the interview guides.

Two research assistants were trained by JM over a period of five days to use a semi-structured interview guide, focus group discussion guide, note-taking templates, consent forms and ethical procedures, including obtaining informed consent from participants. The research assistants conducted key informant interviews and focus group discussions with healthcare professionals and primary caregivers of young children. Data were gathered through semi-structured interviews with relevant key stakeholders, including health and non-health professionals (representatives of community-based organizations) working with young children; and focus group discussions with caregivers of young children. The interviews were conducted in English, Kiswahili or a mix of both languages, based on participants’ preference and level of understanding. The interviews were held in community venues, such as school and church halls within Eastleigh, ensuring privacy throughout. The average duration of the interviews was an hour, while that of the focus groups was two hours.

### Data management and analysis

Interviews and group discussions were audio-recorded, transcribed, and imported into NVivo 20 software for data management and analysis. Framework analysis was used to analyse and interpret the data, following the five stages outlined by Ritchie and Spencer (41): familiarization, developing a coding framework, coding, charting and interpretation. First, the researchers familiarized themselves with the interview data. JM then coded the data against a coding framework that was developed *a priori* from the literature and research question (see S1 Table). Output reports were generated through NVivo, and JM, MT and AA used these to identify themes through an iterative process. The Nurturing Care Framework (42) and the Quality Framework for Parenting Programs (43) were employed in interpreting the findings and their application to the intervention design.

### Ethical procedures

Ethics approval for this study was granted by International Livestock Research Institute (ILRI-IREC2019-15/2), the Kenya National Commission on Science, Technology and Innovation (NACOSTI/P/21/13533), and Nairobi County and Kamukunji sub-County Health Management Team (EOP/NMS/HS/156). Informed consent was obtained from all participants in written form.

## Results

### Participant characteristics

Most of the key informants were female (75%), with a mean age of 42.6 years. They were all Kenyan, had tertiary-level training, and were mainly from the health and nutrition sector, with a few from water and sanitation and community-based organizations. Their roles included advocacy, capacity building, mobilization, coordination, oversight, referral, monitoring and evaluation, and healthcare provision. The focus group discussion participants were all female, with a mean age of 27.7 years, with mostly primary school education (35.3%). Six refugee caregivers participated in the group discussions. Table 1 shows the characteristics of the study participants.

**Table 1:** Characteristics of study participants.

| Characteristics | KIIs<br>*f*(%)<br>n=12 | FGDs<br>*f*(%)<br>n=17 | Total<br>*f*(%)<br>n=29 |
| --- | --- | --- | --- |
| Gender |  |  |  |
| Female | 9 (75) | 17 (100) | 26 (89.7) |
| Male | 3 (25) | - | 3 (10.3) |
| Age |  |  |  |
| Mean | 42.6 | 27.7 | 33.2 |
| SD | 9.29 | 5.16 | 9.99 |
| Highest education level |  |  |  |
| Tertiary (bachelors, diploma, certificate) | 12 (100) | 1 (5.8) | 13 (44.8) |
| Secondary | - | 5 (29.4) | 5 (17.2) |
| Primary | - | 6 (35.3) | 6 (20.7) |
| None | - | 5 (29.4) | 5 (17.2) |
| Refugees | - | 6 (100) | 6 (100) |

The next section provides an analysis of the perceptions of both the key informants and caregivers on current practices in three key areas: health, nutrition and child development. It also highlights barriers and enablers for integration of digital technology within parenting interventions and maps the findings to the intervention design. While this study did not specifically set out to make a comparison between refugees and the host communities, in the few instances where differences between the groups emerged, we highlight them.

### Perceptions on current practices

#### Maternal and child health

Participants stated that antenatal (ANC) and postnatal services (PNC) were widely available through public and private facilities, and accessible to the caregivers in Eastleigh and its surroundings. Public health facilities were preferred by caregivers since their services were free or low cost. While uptake for the first ANC visit was high, only a few caregivers completed the recommended four ANC visits. For PNC, many mothers reportedly missed the first visit typically scheduled two weeks after delivery. Low uptake among the refugees was attributed to lack of awareness about the existence of the services, inflexible vaccination schedules, fear of arrest for lack of legal documentation, and high mobility.

> *“The uptake of first ANC is considerably high. But when it comes to the uptake of the fourth ANC, it’s very low. You’ll find that these… expectant mothers, sometimes, they start…the first ANC late or even don’t complete the four visits*.*” (KII012_CBO representative)*

> *“Like the refugees, maybe they are always on the move and most of them come out at night to seek these services when the facilities, especially the public facilities, are closed to avoid being caught*.*” (KII01_Doctor)*

Participants reported that Community Health Promoters (CHPs), who were essential for outreach efforts, often faced remuneration issues, which led to a lack of motivation and high turnover rates. They also indicated that inadequate CHPs resulted in fewer community health units, which are the basic units for delivering primary healthcare at community level, and thus fewer community members being reached. Additionally, shortage of translators created communication barriers for non-English and non-Swahili speaking patients who were often refugees. Further, poor healthcare worker attitude and scheduling issues contributed to low levels of health seeking and uptake of referrals.

> *“*…*this is now a challenge within our system, when they [refugees] come to the facility, we don’t have translators, language is an issue. And when this mother doesn’t come with somebody who is able to understand Swahili or English, it becomes so difficult. (KII08_Community Health Assistant)*

> *“Yes, except as a human being, you get sometimes the attitude of the healthcare providers becomes a barrier. Yes, maybe the mother is coming in the afternoon, and she is told no, no, no. If you want to attend antenatal [clinic] you come in the morning hours. (KII02_Nutritionist)*

#### Maternal and child nutrition

Breastfeeding initiation at birth was described as high, although adherence to exclusive breastfeeding (EBF) for the first six months was low. The reasons included perceptions of child hunger, mothers needing to resume work, mothers’ inability to produce milk, and lack of interest. Among those not practising EBF, complementary feeding and use of infant formula or other breast milk substitutes, began typically around the second month. Continued breastfeeding up to two years was practised by few caregivers. Participants reported that community sensitization campaigns and the existence of a milk bank in the area had helped increase EBF rates.

> *“Some follow [guidelines from] the clinic, but there are others who say that the children are not satisfied, [and] they give them milk. So, they say they can’t [exclusively] breastfeed until six months [and] must give canned milk*.*” (KII011_CBO representative)*

Refugee communities were perceived to have lower rates of exclusive breastfeeding than the host communities. This was attributed to refugees seldom being reached by sensitization campaigns and the tendency to avoid the formal healthcare system for lack of legal documents.

> *“…there was a time we were dealing with children who are under six [months], and we realized several mothers had already started giving other foods. But these specifically were Somalis, Boranas and Ethiopians… For the locals, most of them were breastfeeding…exclusively…because of the encouragement they were getting from the facility*.*” (KII02-Nutritionist)*

> *“We have…routine sensitization, so nowadays they’re doing it. For six months, they’re breastfeeding, unless the mother is either sick or something, but…there’s a [human] milk bank in Pumwani Maternity*…*” (KII09_Community Health Promoter)*

In both communities, participants reported a preference for foods that were considered to boost milk production such as nourishing soups and porridge. Some dietary differences were noted between refugees and the host community. For instance, the most consumed foods by the refugee mothers, especially those from Somalia, were milk and meat, with limited vegetables and fruits, because of their nomadic pastoralist background. Within the host community, legumes and traditional green vegetables were reported as being predominant.

> *“Lactating mothers, [take] a lot of milk. I think camel milk or something, and then we also have the meat… I don’t see them [consuming vegetables much], but they are so much into pasta, milk and meat…They have also traditional herbs they usually take… periodically, depending on the duration after giving birth*.*” (KII03_Maternal and child health)*

#### Child development

Participants identified positive caregiving behaviours in the community. These included playing with children with or without toys, talking and singing to the children, taking children outdoors from time to time, good hygiene practices to prevent illnesses, and clinic visits for routine growth monitoring and early detection of developmental delays.

> *“For example, you can give the child something to hold. There are other things, like, you can say to the child ‘zingi zingi’, give things, and play with the child. The child will get used to it*.*” (Participant 3, FGD_01)*

However, some harmful practices were identified such as shouting at the child, unregulated screen time and low caregiver involvement in play. Additionally, participation in economic activities was perceived to affect the extent to which caregivers spent time with children.

> *“If you look at how mothers respond to their children, mothers are not playing with them, mothers are not smiling. They just put their children there in the baby walkers and they’re left to watch TV. There are those who are given phones to play with*.*” (KII08_Community Health Assistant)*

> *“Well, you find them now just buying toys…, the other problem is that they don’t play with the children, so they expect these children to know how to play with these toys…they don’t buy toys according to the age of the child, they just buy a toy, the baby will play with the toy*.*” (KII02-Nutritionist)*

Having explored some of the gaps in health, nutrition, and child development encompassing responsive caregiving, play and communication, we explored the barriers and facilitators for digital interventions to further inform the intervention design.

### Barriers and facilitators for digital interventions

The use of digital technology was discussed, first, in the context of access and utilization of smart phones in households within the community, and secondly, the integration of smart phones within parenting interventions. Access to smart phones was seen as limited due to affordability and high insecurity rates within the informal settlements. Cost of internet data was considered a challenge due to low-income levels. However, caregivers had devised strategies for accessing data at low cost, in small chunks, through daily or hourly flash sales from the internet service providers and taking advantage of free Wi-Fi whenever they could. Utilization was often limited by the relatively high illiteracy rates in the informal settlements, language barrier, and low technological know-how.

> *“These caregivers do not have smart phones. I see most or some of the caregivers using [feature phones]. (KII011-CBO representative)*

> *“You can’t be online for many hours. Perhaps after you’ve worked till like 3:00 pm, you buy a 1-hour bundle and go online. Once that 1-hour is over, you go offline till the next day*.*” (Participant, FGD_02)*

> *“…how do I put it, not really ignorance but language barrier, yeah, so if we can bridge that then I don’t think they will have any issue using their phone [for you] to be able to relay the messages. (KII03-Maternal and child health)*

The use of smart phones in health, nutrition, and parenting interventions was perceived to be limited in the Eastleigh context. Even among the interventions that used phones, this was limited to only short text messages through feature phones. Most caregivers indicated that they had never used their phones for any health, nutrition or parenting-related activities.

> *“There is not any other service that is delivered through smartphones*.*” (KII03-Maternal and child health)*

> *Not really, this is just a one-on-one kind of messaging or service provision. At the facility or the community level, we don’t have cases where we are using phones or smart phones*.*” (KII07_Community Health Strategy)*.

Health workers and caregivers strongly recommended the integration of technology into interventions, which was considered beneficial, especially to refugee communities who are often left out of service delivery. Other considerations included supporting smart phone ownership and being cognizant of intra- or inter-family sharing, translating content to refugee-spoken languages, and addressing caregiver illiteracy by providing an audio option. There was a keen interest in interventions that encouraged health seeking behaviour, for instance, through CHPs or technology, such as sending out reminders for ANC and PNC visits.

> *“The moment those you are targeting are taught even for one day, or they use that app [the digital intervention] …, it will encourage them. They will learn a lot. (Participant 7, FGD_01)*

> *“And my recommendation…having the information through your mobile phone is very important because you can easily access it with a lot of ease…” (KII012_CBO representative)*

> *“You also need to see which group you are targeting, whether they have the phones, or they share within one household. (KII06_Water Sanitation and Hygiene)*

### Application of findings to intervention design

The findings were useful in informing the intervention design, including aligning the content with community needs, identifying the delivery mode, and workforce. This process followed the Quality Framework for Parenting Programs (43). Table 2 shows the areas in which the intervention design was informed by the findings based on the framework.

**Table 2:** Mapping findings to intervention design decisions.

| Component | Findings | Application in intervention design |
| --- | --- | --- |
| Intervention content | ANC and PNC services not optimally utilized.<br>Poor dietary diversity.<br>Some caregivers showed inappropriate caregiving practices such as shouting at child.<br>Hardly any interventions targeting health and caregiving behaviour change. | Prompting to encourage certain behaviours e.g. reminders on immunization dates, health seeking, caregiver-child play and communication, nutritious food options.<br>Self-monitoring through recording practices e.g. number of times a child breastfeeds; foods consumed in 24-hour period.<br>Personalized messaging and feedback to encourage positive behaviour.<br>These three strategies have been shown to be effective in behaviour change (44, 45). |
| Delivery modality | Know-how and access to smart phones.<br>Preference for interactive communication.<br>Low literacy.<br>Cost of data bundles hinders internet access and could limit application use. | An interactive/two-way communication was adopted. The mobile application had a component for messaging the caregivers on the identified topics, tracking progress based on caregiver-inputted data and providing feedback based on the inputted data.<br>An audio option provided in addition to the written messages.<br>Train caregivers and CHPs on the intervention application and smart phone use.<br>Provide smart phones and data bundles to caregivers and CHPs for the feasibility study. |
| Workforce and resources | Existing resources include private and public healthcare providers, and community health promoters (CHP)s.<br>CHPs poorly compensated and facilitated to carry out their work.<br>Poor awareness among some of the community members, and refugees, about existing healthcare services. | Use existing resources such as CHPs.<br>Compensate the CHPs for their time and expenses in implementing the intervention.<br>Identify health facilities and services offered, including maternity, health centres and milk banks, for referral of caregivers in the study. |
| Context | • Refugee caregivers and their children often left out of health interventions.<br>• Both refugee and host community often face similar challenges. | • Target primary caregivers of young children, 0-3 years.<br>• Target both the refugees and host communities to avoid stigma and build goodwill within the community. |

## Discussion

This study explored community perceptions on current health, nutrition, caregiving and child development practices among urban refugees and host community in a challenging context. We identified barriers and facilitators for the adoption of digital parenting interventions. We further mapped the findings against different intervention decision points. While there were some perceived positive practices among refugee and host communities, they were described as insufficient, especially around breastfeeding, dietary diversity, ANC, PNC, and positive parenting. Additionally, technological interventions were still underutilized despite an increase in access to smart phones as participants indicated little to no knowledge of any such interventions at the time of the study.

Caregiver knowledge and skills are critical because of their influence on child development outcomes (46). Research has linked caregiver knowledge of early child development to caregiving competence and shown that knowledge of child development moderates the impact of self-efficacy on parenting competence (47). Further, caregiver knowledge, skills, and literacy have positive effects on the health and nutrition of family members, including young children (23, 48). Our study found that refugees were perceived to have lower levels of caregiving knowledge. However, it is important to note that this was simply a perception among participants and likely to have as much to do with prejudice as reality. There is weak evidence that refugees have lower levels of caregiving or parenting knowledge compared to the host community (46).

The perceived high level of awareness about maternal and child health and nutrition within the host community was possibly due to community health outreach programmes. Kenya has experienced several community-led sensitization campaigns on Maternal, Newborn, and Child Health (MNCH) in recent years, most notably the ‘Linda Mama’ initiative, which provides free ANC, delivery and PNC (17). Both community-based and facility-based healthcare services had limited reach among refugees,due to factors such as language barriers and lack of legal documentation, further marginalizing them. Other studies found similar barriers to healthcare access by refugees, including insufficient knowledge on how to access healthcare services in the host country, in addition to financial constraints and language barriers (49). There was also perceived discrimination and a poor attitude by healthcare staff discouraging refugees from seeking healthcare services (49), which was also true of participants in our study.

Caregiver involvement in play and communication is crucial for their bonding with the child (50). However, economic participation, likely in informal jobs (51), may hinder caregiver involvement. These jobs reduced the amount of time caregivers could spend on caregiving as did the number of children. Our finding supports previous research which indicates that mothers working in the informal sector often lacked the time to engage in play-based learning with their children (52, 53). Sensitization on the importance of play and communication in the development of the child with particular emphasis on the use of low- or no-cost locally available play materials would be crucial in enhancing child development in the community. The sensitization could be coupled with economic empowerment programs to address livelihood challenges and help safeguard at-risk communities. Further, establishment of safe outdoor play spaces within the communities, and well-equipped play areas in health facilities could serve to promote play.

There was a rise in adult smart phone ownership in Kenya from 54% to 61% between June 2022 and June 2023 (54). Despite this, we found little evidence of the use of phones to aid parenting, except for gaming or watching cartoons. While mHealth interventions are not without drawbacks, (29), there remains great potential for the utilization of phones in interventions aimed at enhancing health, nutrition and development of young children if barriers are addressed. Evidence on the effectiveness of mHealth interventions is well documented (24). Interventions such as those encouraging EBF, health seeking, ANC, PNC, immunization and dietary diversity, would do well to explore integration of smart phone technology. Any interventions looking to use smart phones should be mindful of phone access, internet costs, and ensure adequate training of caregivers and CHPs. Sim cards could be registered through the project for refugee inclusion, granted this needs to be negotiated with the telecommunications companies.

### Strengths and limitations

This study had many strengths, including having a diverse range of participants from the health system and the community, ensuring a variety of perspectives and experiences. However, there was low participation of refugees for two reasons. First, they are not typically included in the public health sector as staff, hence their exclusion from the key informant interviews. Secondly, the study was only conducted in English and Kiswahili which are the main languages spoken in the context but may not have been readily understood by some of the refugees. However, we believe that this number was representative as two thirds of the refugee caregivers contacted participated. Finally, while strategies to reduce bias such as using a pre-defined coding framework were employed, there may still have been researcher bias in the interpretation of the findings as is inevitable in qualitative studies.

## Conclusion

Both refugee and host communities in urban informal settlements experience unique challenges in seeking and accessing maternal health, nutrition and child development services, with some challenges being unique to the embedded refugees. While healthcare providers work to bridge the gap, workforce shortfalls and other resource gaps continue to hinder access. Underutilization of technology, despite advances in mHealth, points to the critical need to prioritize its integration within maternal and child health initiatives, focusing on nutrition and child development. Embracing digital solutions could enhance outcomes for caregivers and children. The insights from this study offer important considerations in designing effective, contextually appropriate, digital parenting interventions to ensure acceptance within the community.

## Data Availability

All relevant data are within the manuscript and its Supporting Information files.

## Acknowledgement

The authors would like to thank the Kamukunji Sub-County Health Management Community for providing an entry into the community. We also thank the enumerators: Barack Aoko and Kevin Wekesa, and the Community Health Promoters: Samira Hussein and Eunice Inanga. We are grateful to all the caregivers and healthcare workers who took part in this study.

## Supporting information

**S1 Table. Coding framework**

## References

1. Ritchie H, Mathieu E. How many people die and how many are born each year? Our world in data 2023.

2. Trends in maternal mortality 2000 to 2020: Estimates by WHO, UNICEF, UNFPA, World Bank Group and UNDESA/Population Division. Geneva: WHO; 2023.

3. UNICEF, WHO, World Bank Group Joint Malnutrition Estimates, May 2023 Edition. Global prevalence and numbers affected 2000-2022.: UNICEF, World Health Organization, World Bank; 2023.

4. Transforming our world: The 2030 agenda for sustainable development. 2015.

5. World urbanization prospects. United Nations; 2014.

6. Baeumler A, D’Aoust O, Das MB, Gapihan A, Goga S, Lakovits C, et al. Demoraphic trends and urbanization. Washington DC: World Bank; 2021.

7. World Cities Report 2024: Cities and climate action. UN-HABITAT; 2024.

8. Global trends: Forced dispacement in 2022. Copenhagen: UNHCR; 2023.

9. A study on health vulnerabilities of urban migrants in the greater Nairobi. Nairobi: IOM; 2013.

10. Gardner J, Lenette C, Al Kalmashi R. “Who is the host? Interrogating hosting from refugee-background women’s perspectives.”. Journal of Intercultural Studies. 2022;43(5):621–38.

11. Kabue M, Abubakar A, Ssewanyana D, Angwenyi V, Marangu J, Njoroge E, et al. A community engagement approach for an integrated early childhood development intervention: A case study of an urban informal settlement with Kenyans and embedded refugees. BMC Public Health. 2022;22:711.

12. Pega F, Pabayo R, Benny C, Lee E-Y E-Y, Lhachimi S, Liu S. Unconditional cash transfers for reducing poverty and vulnerabilities: effect on use of health services and health outcomes in low-and middle-income countries. Cochrane Database of Systematic Reviews. 2022(3):CD011135.

13. Ochola S, Ogada IA, Odera CA. Predictors of the amount of intake of ready-to-use therapeutic foods among children in outpatient therapeutic programs in Nairobi, Kenya. Food Science & Nutrition. 2022;10(4):1135–45.

14. Njoroge MN, Munene F. Nutrition survey conducted in the slums of Nairobi County. Nairobi: Concern Worldwide-Kenya; 2017.

15. Mberu BU, Haregu TN, Kyobutungi C, Ezeh AC. Health and health-related indicators in slum, rural, and urban communities: A comparative analysis. Global Health Action 2016;9:33163.

16. Hone T, Goncalves J, Seferidi P, Moreno-Serra R, Rocha R, Gupta I, et al. Progress towards universal health coverage and inequalities in infant mortality: an analysis of 4.1 million births from 60 low-income and middle-income countries between 2000 and 2019. The Lancet Global Health. 2019;12(5):e744–e55.

17. Orangi S, Kairu A, Ondera J, Mbuthia B, Koduah A, Oyugi B, et al. Examining the implementation of the Linda Mama free maternity program in Kenya. The International Journal of Health Planning and Management. 2021;36(6):2277–96.

18. Fan YW, Fan HSL, Shing JSY, Ip HL, Fong DYT, Lok KYW. Impact of baby-friendly hospital initiatives on breastfeeding outcomes: Systematic review and meta-analysis. Women and Birth. 2025;38(2):101881.

19. Daily B. Affordable housing lessons for Kenya from Colombia. 2019.

20. Little MT, Roelen K, Lange BCL, Steinert JI, Yakubovich AR, Cluver L, et al. Effectiveness of csh-plus programmes on early childhood outcomes compared to cash transfers alone: A systematic review and meta-analysis in low-and middle-income countries. PLoS medicine. 2021;18(9):e1003698.

21. Knop MR, Nagashima-Hayashi M, Lin R, Saing CH, Ung M, Oy S, et al. Impact of mHealth interventions on maternal, newborn, and child health from conception to 24 months postpartum in low- and middle-income countries: a systematic review. BMC Medicine. 2024;22(1):196.

22. De P, Pradhan MR. Effectiveness of mobile technology and utilization of maternal and neonatal healthcare in low and middle-income countries (LMICs): a systematic review. BMC Women’s Health. 2023;23(1):664.

23. DeWalt DA, Hink A. Health literacy and child health outcomes: a systematic review of the literature. Pediatrics. 2009;124(Supplement_3):S265–S74.

24. Colaci D, Chaudhri S, Vasan A. mHealth interventions in low-income countries to address maternal health: A systematic review. Annals of Global Health. 2016;82(5):922–35.

25. Sondaal SFV, Browne JL, Amoakoh-Coleman M, Borgstein A, Miltenburg AS, Verwijs M, et al. Assessing the effect of mHealth interventions in improving maternal and neonatal care in low-and middle-income countries: A systematic review. PloS One. 2016;11(5):e0154664.

26. Carmichael SL, Mehta K, Srikantiah S, Mahapatra T, Chaudhuri I, Balakrishnan R, et al. Use of mobile technology by frontline health workers to promote reproductive, maternal, newborn and child health and nutrition: A cluster randomized controlled trial in Bihar, India. Journal of Global Health. 2019;9(2).

27. Feroz A, Jabeen R, Saleem S. Using mobile phones to improve community health workers’ performance in low-and-middle income countries. BMC Public Helath. 2020;20(49).

28. Tomlinson M, Rotheram-Borus MJ, Doherty T, Swendeman D, Tsai AC, Ijumba P, et al. Value of a mobile information system to improve quality of care by community health workers South African Journal of Information Management. 2013;15(1).

29. Ikwunne T, Hederman L, Wall PJ. Designing mobile health for user engagement: The importance of socio-technical approach arXiv preprint arXiv. 2021:2108.09786.

30. Wallerstein N, Bonnie D, Oetzel JG, Minkler M, eds. Community-based participatory research for health: Advancing social and health equity: John Wiley & Sons; 2017.

31. Woolf SH, Zimmerman E, Haley A, Krist AH. Authentic engagment of patients and communities can transform research, practice, and policy. Health Affairs. 2016;35(4):590–4.

32. Mac-Garity-Palmer R, Saw A, Keys CB. Community engagment in psychosocial interventions with refugees from Asia: a systematic review. Asian American Journal of Psychology. 2023;14(2):117–32.

33. Hacker K, Tendulkar SA, Rideout C, Bhuiya N, Trinh-Shevrin C, Savage CP, et al. Community capacity building and sustainability: outcomes of community-based participatory research. Progress in Community Health Partnerships: Research, Education, and Action. 2012;6(3):349.

34. Cunningham-Erves J, Barajas C, Mayo-Gamble TL, McAfee CR, Hull PC, Sanderson M, et al. Formative research to design a culturally-appropriate cancer clinical trial education program to increase participation of African American and Latino communities. BMC Public Health. 2020;20:840.

35. Statistics KNBo. 2019 Kenya population and housing census. Nairobi: Kenya National Bureau of Statistics; 2019.

36. Asoka GWN, Thuo ADM, Bunyasi MM. Effects of Population Growth on Urban Infrastructure and Services: A Case of Eastleigh Neighborhood Nairobi, Kenya. Journal of Anthropology & Archaeology. 2013:41–56.

37. Nyamasege CK, Kimani-Murage EW, Wanjohi M, Kaindi DWM, Wagatsuma Y. Effect of maternal nutritional education and counselling on children’s stunting prevalence in urban informal settlements in Nairobi, Kenya. Public health nutrition. 2021;24(12):3740–52.

38. Kithinji S, Mambo S, Kyallo F. Factors influencing malnutrition among children aged 6-59 months in Kamukunji Sub-County, Nairobi County, Kenya. East African Medical Journal. 2024;101(12):7648–56.

39. Sudfeld CR, McCoy DC, Fink G, Muhihi A, Bellinger DC, Masanja H, et al. Malnutrition and its determinants are associated with suboptimal cognitive, communication, and motor development in Tanzanian children. The Journal of nutrition. 2015;145(12):2705–14.

40. Suri H. Purposeful sampling in qualitative research synthesis. Qualitative Research Journal. 2011;11(2):63–75.

41. Ritchie J, Spencer L. Qualitative data analysis for applied policy research. In: Bryman A, Burgess RG, editors. Analyzing qualitative data. London & New York: Routledge; 2002. p. 173–94.

42. Nurturing care for early childhood development: a framework for helping children survive and thrive to transform health and human potential Geneva: World Health Organization; 2018.

43. Bozmamadova, M, Laouali R, Sosa Walde P. Guidance Note on Designing and Implementing Early Childhood Parenting Programs. 2025:1–20.

44. EUFIC. Behaviour change models and strategies. 2014.

45. Van Achterberg T, Huisman-De Waal GGJ, Ketelaar NABM, Oostendorp RA, Jacobs JE, Wollersheim HCH. How to promote healthy behaviours in patients? An. Health promotion international. 2011;26(2):148–62.

46. Rowe ML, Denmark N, Harden BJ, Stapleton LM. The role of parent education and parenting knowledge in children’s language and literacy skills among White, Black and Latino families. Infant and Child Development. 2016;25(2):198–220.

47. Hess CR, Teti DM, Hussey-Gardner B. Self-efficacy and parenting of high-risk infants: The moderating role of parnt knowledge of infant development. Journal of Applied Development. 2004;25(4):423–37.

48. de Buhr E, Tannen A. Parental health litearcy and health knowledge, behaviours and outcomes in children: a cross-sectional survey. BMC Public Health. 2020;20:1–9.

49. Mangrio E, Sjögren Forss K. Refugees’ experiences of healthcare in the host country: a scoping review. BMC Health Services Research. 2017;17:1–16.

50. Ginsburg KR, Communications Co, Child CoPAo, Health F. The importance of play in promoting healthy child development and maintaining strong parent-child bonds. Pediatrics. 2007;119(1):182–91.

51. Golla AM. Engaging informal women entrepreneurs in East Africa: approaches to greater formality. An ILO-WED Issue Brief. 2017.

52. Okelo K, Nampijja M, Ilboudo P, Muendo R, Oloo L, Muyingo S, et al. Evaluating the effectiveness of the Kidogo model in empowering women and strengthening their capacities to engage in paid labor opportunities through the provision of quality childcare: a study protocol for an exploratory study in Nakuru County, Kenya. Humanities and Social Sciences Communications. 2022;9(1):1–7.

53. Horwood C, Hinton R, Haskins L, Luthuli S, Mapumulo S, Rollins N. ‘I can no longer do my work like I used to’: a mixed methods longitudinal cohort study exploring how informal working mothers balance the requirements of livelihood and safe childcare in South Africa. BMC Women’s Health. 2021;21:1–15.

54. Obura F. Smartphone Penetration in Kenya Rises 61 per cent to 30.8Million. The Kenyan Wall Street; 2023.

